# Development of a sensitive trial-ready poly(GP) CSF biomarker assay for *C9orf72*-associated frontotemporal dementia and amyotrophic lateral sclerosis

**DOI:** 10.1101/2021.12.14.21267456

**Authors:** Katherine M Wilson, Eszter Katona, Idoia Glaria, Imogen J. Swift, Aitana Sogorb-Esteve, Carolin Heller, Arabella Bouzigues, Amanda J Heslegrave, Saurabh Patil, Susovan Mohapatra, Yuanjing Liu, Jaya Goyal, Raquel Sanchez-Valle, Robert Laforce, Matthis Synofzik, James B. Rowe, Elizabeth Finger, Rik Vandenberghe, Chris R. Butler, Alexander Gerhard, John van Swieten, Harro Seelaar, Barbara Borroni, Daniela Galimberti, Alexandre de Mendonça, Mario Masellis, Carmela Tartaglia, Markus Otto, Caroline Graff, Simon Ducharme, Andrea Malaspina, Henrik Zetterberg, Ramakrishna Boyanapalli, Jonathan D Rohrer, Adrian M Isaacs, on behalf of the Genetic FTD Initiative (GENFI)

## Abstract

A GGGGCC repeat expansion in the *C9orf72* gene is the most common cause of genetic frontotemporal dementia (FTD) and amyotrophic lateral sclerosis (ALS). As potential therapies targeting the repeat expansion are now entering clinical trials, sensitive biomarker assays of target engagement are urgently required. We utilised the single molecule array (Simoa) platform to develop an immunoassay for measuring poly(GP) dipeptide repeat proteins (DPRs) generated by the repeat expansion in CSF of people with *C9orf72*-associated FTD/ALS. We show the assay to be highly sensitive and robust, passing extensive qualification criteria including low intra- and inter-plate variability, a high precision and accuracy in measuring both calibrators and samples, dilutional parallelism, tolerance to sample and standard freeze-thaw and no haemoglobin interference. We used this assay to measure poly(GP) DPRs in the CSF of samples collected through the Genetic FTD Initiative. We found it had 100% specificity and 100% sensitivity and a large window for detecting target engagement, as the *C9orf72* CSF sample with the lowest poly(GP) signal had 8-fold higher signal than controls and on average values from *C9orf72* samples were 38-fold higher than controls, which all fell below the lower limit of quantification of the assay. These data indicate that a Simoa-based poly(GP) DPR assay is suitable for use in clinical trials to determine target engagement of therapeutics aimed at reducing *C9orf72* repeat-containing transcripts.

## Introduction

A GGGGCC repeat expansion in the first intron of *C9orf72* is the most common genetic cause of both amyotrophic lateral sclerosis (ALS) and frontotemporal dementia (FTD) accounting for 38% and 25% of familial cases respectively^1^. Healthy individuals most commonly have two repeats^2^, whilst people with a *C9orf72* repeat expansion (C9FTD/ALS) can carry hundreds to thousands of repeats^3,4^. The repeats are transcribed in both sense and antisense direction, leading to the formation of RNA aggregates termed RNA foci^5–8^. In addition, repeat-associated non-ATG (RAN) translation of the repeat expansion leads to the production of dipeptide repeat proteins (DPRs). Translation occurs in all three frames from both sense and antisense transcripts producing five different dipeptide species, poly(GA), poly(GP), poly(GR), poly(PR) and poly(PA). Therapies targeting the *C9orf72* repeat expansion such as small molecules^9,10^, antisense oligonucleotides (ASOs)^8,11–16^, siRNAs^17^, microRNAs^18^ and CRISPR-based approaches^19–21^ are rapidly being developed. ASOs targeting the repeat expansion or *C9orf72* transcripts have been shown to reduce both RNA foci and DPR levels in human iPSC-neurons^11,12,15^ and *C9orf72* mouse models^8,13–16^. In order to progress therapies from the bench to the bedside, biomarkers of disease that can reflect target engagement are needed. An important breakthrough was the discovery that poly(GP) can be detected in the cerebrospinal fluid (CSF) of people with C9FTD/ALS using Meso Scale Discovery (MSD) immunoassays, indicating its potential as a target engagement biomarker^15,22^. Levels of poly(GP) in CSF were not found to correlate with clinical disease markers or neurofilament CSF levels, a non-disease specific biomarker of neurodegeneration^15,22^. Encouragingly, ASO treatment of mice models has been shown to lead to durable, decreased poly(GP) levels both in brain tissues and mouse CSF, showing that CSF poly(GP) levels could be used as a pharmacodynamic biomarker^14–16^.

The single molecule array (Simoa) platform measures immuno-complexes bound to microscopic beads that are isolated in arrays of microwells, large enough for a single bead. Using digital detection and Poisson distribution for quantification, the Simoa platform enables single molecule detection^23^. Here, we developed a sensitive, qualified poly(GP) assay using Simoa technology. Following extensive assay development and qualification we measured poly(GP) levels in CSF collected through the Genetic FTD Initiative (GENFI). We found a clear, 8-fold difference in signal between controls and the C9FTD patient with the lowest measured poly(GP) levels. Using this cohort, the assay had sensitivity of 100% and specificity of 100%. Despite assay sensitivity, no correlations between poly(GP) and clinical features were found. The assay was further optimised for plasma however, matched donor plasma samples were found to have poly(GP) levels below the lower limit of detection (LLOQ).

## Materials and Methods

### Participants

Fifty-five participants were recruited from the Genetic FTD Initiative (GENFI), a natural history study of genetic FTD based across 27 sites in Europe and Canada^24^. Participants included 15 symptomatic *C9orf72* expansion carriers (14 with behavioural variant FTD (bvFTD) and 1 with ALS), 25 presymptomatic C9orf72 expansion carriers and 15 non-carrier relatives, as controls. Pathogenic *C9orf72* expansion length was defined as more than 30 repeats. Participants consisted of 23 men and 32 women, with a mean (standard deviation) age of 49.4 (13.9) years old at sample collection. Within the disease groups: presymptomatic *C9orf72* expansion carriers, 11 men and 14 women, 41.0 (10) years old and symptomatic *C9orf72* expansion carriers, 10 men and 5 women, 64.7 (8.5) years old. 15 healthy controls were recruited over the same time period: 2 men and 13 women, 48. 2 (11.2) years old. All people in the study underwent a clinical assessment consisting of a medical history with the participant and informant, and physical examination, with symptomatic status diagnosed by a clinician who was an expert in the FTD field^25–29^. All participants also underwent 3D T1-weighted magnetic resonance imaging of the brain. Volumetric measures of whole brain and cortical regions were calculated using a previously described method that uses the geodesic information flow (GIF) algorithm, which is based on atlas propagation and label fusion^30^.

The study procedures were approved by local ethics committees at each of the participating sites and participants provided informed written consent.

### CSF and plasma collection

CSF and plasma were collected, processed and stored in aliquots at −80°C according to standardised procedures^31^.

### NfL plasma assay

Plasma NfL concentration was measured in 8 matched symptomatic CSF donors, 10 matched presymptomatic CSF donors and 5 matched healthy control CSF donors using the multiplex Neurology 4-Plex A kit (102153, Quanterix, Billerica, MA) on the Simoa HD-1 Analyzer following manufacturer’s instructions.

### Antibodies

Rabbit Polyclonal antibodies ‘GP57’ and ‘GP60’ were produced using a synthetic polypeptide, GP(32) as antigen and provided by Wave Life Sciences. An alternative polyclonal anti-GP antibody ‘GP6834’ was custom-made by Eurogentec, using GP(8) as antigen. The monoclonal poly(GP) antibody TALS 828.179 was obtained from the Developmental Studies Hybridoma Bank, deposited by Target ALS Foundation. Antibody details are summarised in Table 1.

**Table 1.** Details of polyclonal and monoclonal antibodies tested in Simoa poly(GP) assays. Rabbit polyclonal antibodies were affinity purified prior to biotinylation and testing.

| Anti-GP antibody name | Peptide used as antigen | Monoclonal /Polyclonal | Source |
| --- | --- | --- | --- |
| GP57 | GP(32) | Rabbit polyclonal | Custom made |
| GP60 | GP(32) | Rabbit polyclonal | Custom made |
| GP6834 | GP(8) | Rabbit polyclonal | Custom made |
| mGP | GP(8) | Mouse monoclonal | TALS 828.179 |

Antibody bead conjugation and biotinylation was performed as recommended by Quanterix’s Homebrew Assay Development guide. Briefly, 0.3 ml of carboxylated paramagnetic beads were conjugated with 0.2 mg/ml antibody and 0.3 mg/ml 1-Ethyl-3-(3-dimethylaminopropyl) carbodiimide (EDC) with conjugation performed at 2-8°C. This required 80 µg of input antibody. For each biotinylation, 130 µg of antibody was used at 1 mg/ml and a 40:1 ratio of NHS-PEG_4_-biotin to antibody.

### Assay Optimisation

Optimisation of the poly(GP) Simoa assay was performed by testing: 2 step vs 3 step assay design, detector antibody concentrations from 0.3 µg/ml to 1.5 µg/ml, SGB concentrations from 50 pM to 150 pM, the inclusion of helper beads at different ratios or not at all. Multiple assay combinations were run in parallel to enable selection of optimal conditions. GST-GP32 standard curve was prepared from 2 starting stocks (15000 pg/ml and 1500 pg/ml), serially diluting down from both in diluent A (Quanterix) to create a 9-point standard curve + blank. High (140 pg/ml), middle (75 pg/ml) and low (15 pg/ml) quality control (QC) samples were prepared independently for each assay from a 1500 pg/ml stock of GST-GP32. A positive control human CSF sample from *C9orf72* expansion carriers (QC4) was created by pooling a small volume of CSF from the 40 *C9orf72* expansion carriers in the GENFI cohort.

### Curve fitting

To establish best curve fitting we followed a previously described workflow^32^. Firstly, heteroscedasticity (the unequal variability of a variable across a range of values of a second variable that predicts it) was assessed plotting the standard deviation of the average number of enzyme labels per bead (AEB) signals from the calibrators from seven assays, against their concentration (Figure S1A). As the data showed heteroscedasticity, weighting was determined by plotting log(SD of signals) against log(mean of signals) (Figure S1B). After applying linear regression and determining the slope value (k), weighting was then calculated using the following formula: Weighting = 1/Y^2k^ = 1.9474. Curves were recalculated using four parameter logistic (4PL) and five parameter logistic (5PL), with no weighting, 1.9474, or 2 weighting. Curve fits were assessed using criteria that relative errors (RE) and CV for calibrators were +/- 15%, and RE and CV for anchor points (1 pg/ml) were +/- 20%. Curve fitting with 4PL 1/Y^2^ was selected as it led to all calibrator points passing these criteria (Figure S1C).

### Poly(GP) Simoaassay

The optimised Simoa assay (performed on an HD-X instrument, which is an upgraded version of the HD-1 instrument) using TALS 828.179 monoclonal antibody (mGP) beads as capture and a combination of biotinylated GP57 and GP60 (termed GP57*-60*) as detector used the following assay conditions: 2 step assay, 0.3 µg/ml detector antibody (GP57*-60*), 50 pM streptavidin-β-D-galactosidase (SBG), 150000 assay beads (mGP) with 350000 helper beads. CSF was thawed on ice and diluted 1:2 with diluent A (Quanterix). 250 µl per sample was loaded into the sample plate to allow for duplicate measures. Analysts were blind to clinical and genetic status of samples.

Plasma samples were thawed on ice, prior to centrifugation at 14000 rcf for 15 minutes at room temperature. 125 µl of plasma was then diluted with 125 µl of lysate diluent B (Quanterix) to allow duplicate measures per sample. Standard curve was prepared in lysate diluent B diluted 1:2 with control human plasma. Analysts were blind to genetic status of samples.

### Statistical analysis

Statistical analysis was carried out using GraphPad Prism software. Data was tested for normality prior to appropriate parametric or non-parametric tests. Mann-Whitney tests were used for comparing two groups, for more than two groups Kruskal-Wallis tests and Dunn’s multiple comparisons test were used. To assess correlations between poly(GP) and clinical features Spearman rho and p (two-tailed) values were calculated.

## Results

### Development of poly(GP) Simoa assay

To develop a sensitive poly(GP) Simoa assay we first optimised assays using the Simoa HD-1 analyser. We tested a mouse monoclonal anti-GP antibody (mGP) and a range of affinity purified rabbit polyclonal antibodies (GP57, GP60 and GP6834) raised against different length GP peptides (Table 1). As the long-term goal was to have sufficient antibody quantities for use in a biomarker assay in clinical trials, we combined antibodies GP57 and GP60, which were both raised against a GP32 peptide. We found that using the monoclonal antibody as capture and the combined polyclonal antibodies as detector gave the highest signal to noise ratios for the calibrators and lowest lower limit of quantification (LLOQ) for measurement of a GST-GP32 standard peptide (Figure 1). While use of mGP for both capture and detection would have been preferable, due to unlimited supply, even after assay optimisation the mGP + mGP* assay (where * indicates the biotinylated detector antibody) was over 10-fold less sensitive (LLOQ 15.8 pg/ml) than mGP + GP57*-60* (LLOQ 1.04 pg/ml) (Figure 1). As mGP + GP57*-60* showed the highest sensitivity, we took this assay forward. To ensure compatibility in the long-term, we next transferred the assay to the newer Simoa HD-X platform. We found the assay required re-optimisation, with the greatest benefit gained from changing the standard curve diluent from lysate diluent B (HD-1) to diluent A (HD-X) (Figure 2A). In addition, SBG was lowered from 100 pM to 50 pM for the final HD-X assay, with an LLOQ of 1.17 pg/ml (Figure 2B).

**Figure 1.**
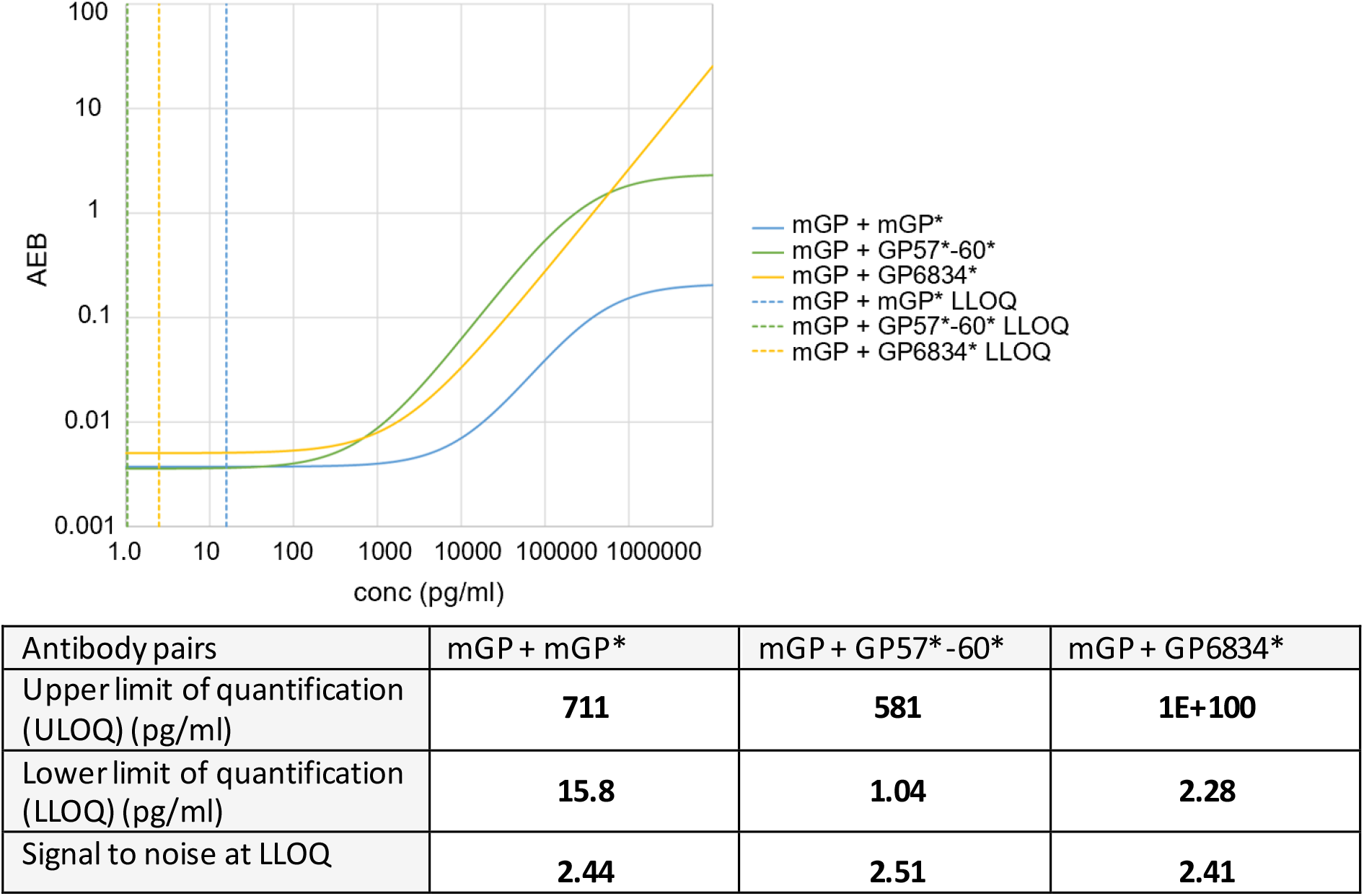
Comparison of monoclonal and polyclonal anti-poly(GP) antibodies in Simoa homebrew assays. Homebrew Simoa assay conditions were optimised using different capture antibodies and detector antibodies (^*^). mGP = monoclonal poly(GP) antibody (TALS 828.179). GP57*-60^*^ is a combination of two custom polyclonal antibodies ‘GP57’ and ‘GP60’. GP6834 is an alternative custom made poly(GP) antibody. Dashed lines show predicted LLOQs for each optimised assay respectively (mGP + mGP^*^, mGP + GP57^*^-60^*^, mGP + GP6834^*^), calculated using Quanterix assay developer tool, after running 6-point standard curves using GST-GP32 as standard.

**Figure 2.**
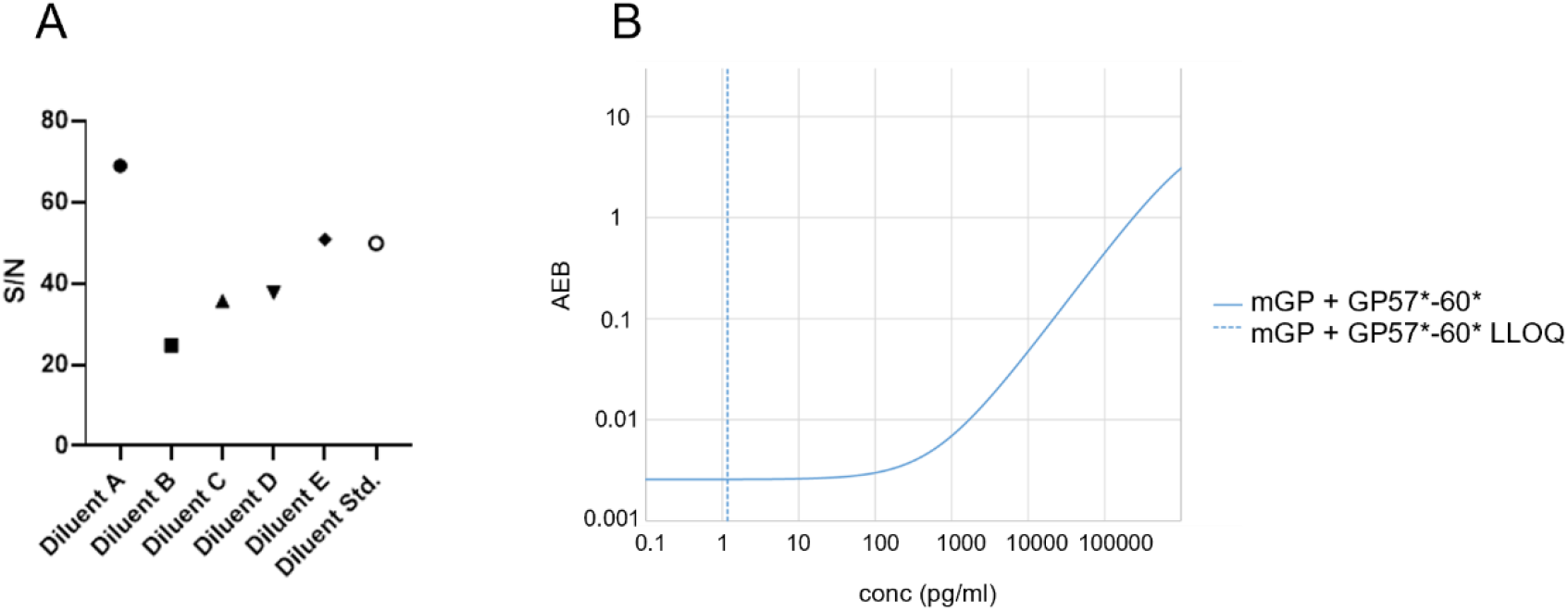
Transfer of poly(GP) assay onto Simoa HD-X. **A)** Effect of sample diluents was assessed by comparing signal/noise (S/N) using control human CSF spiked with 25 pg/ml GST-GP32 standard, diluted 1 in 2 with different Quanterix diluents. Samples were run in duplicate on a single 2 step Simoa assay (HD-X), using mGP + GP57^*^-60^*^ Homebrew assay. **B)** Standard curve produced from optimised mGP + GP57^*^-60^*^ HD-X Simoa assay, using GST-GP32 as standard. LLOQ at 1.17 pg/ml shown by dashed line.

### Qualification of Simoa poly(GP) assay

To prepare this assay for use in clinical trials it was evaluated using standard biomarker assay qualification criteria (Table 2). Precision performance was assessed by analysing standard curves from 7 independent assays, performed by two independent researchers. Coefficient of variation (CV) was <20% for all standard curve points (Figure 3A and Table S1). Difference from total (DFT) (difference between predicted and actual concentration of calibrators) was below 20% for all calibrators in 6/7 assays (Figure 3B and Table S2). LLOQ was identified as 1 pg/ml with upper limit of quantification at 200 pg/ml. Quality control (QC) samples were prepared by spiking the standard reference material GST-GP32 into diluent A. Upper QC (150 pg/ml), Middle QC (75 pg/ml) and Lower QC (5 pg/ml) all showed CVs <20% after 7 independent runs (Figure 3C and Table S3). DFTs were below 25% for QCs in 7 assay runs (Figure 3E and Table S4). Intraplate variability was assessed by measuring 3 sets of QCs across a plate within a single assay, with CV <5% for all 3 QCs (Figure 3F and Table S5). An endogenous matrix QC sample (QC4) was generated by pooling human CSF from *C9orf72* expansion positive donors. Poly(GP) concentration of QC4 was measured in 4 independent assays and the CV was <20% (Figure 3G and Table S7). Intermediate precision was further tested by measurement of QC samples prepared 3 times. This was repeated by a second analyst (Figure 3D and Table S6). CV was <20% for the sets of QCs prepared independently and between the two analysts.

**Table 2.** Biomarker assay qualification criteria for poly(GP) Simoa assay. Coefficient of variation (CV) = (Standard Deviation / Mean) ^*^ 100. Difference from Total (DFT) = difference from predicted concentration of calibrators (pg/ml) from actual, as % of actual. Quality Control samples (QCs) were prepared using GST-GP32 in diluent A.

| Parameter | Criteria | Data |
| --- | --- | --- |
| Precision and accuracy measuring calibrators | 75% of calibrators CV ≤ 20% and 75% of calibrators DFT $\leq \pm$ 20%. | Figure 3A, 3B. Supplementary table 1 and table 2. |
| Precision and accuracy measuring QC samples | High (140 pg/ml), Medium (75 pg/ml) and Low (15 pg/ml) QCs CV ≤ 20% and DFT $\leq \pm$ 20%. | Figure 3C, 3E. Supplementary table 3 and 4. |
| Intra- and interplate reproducibility | Repeat measure of QC samples across multiple plates and positioned across a single plate CV ≤ 20%.<br>3 sets QC samples prepared independently, in two independent assays by two analysts, CV ≤ 20% and DFT $\leq \pm$ 20%. | Figure 3D, 3F. Supplementary table 5 and 6. |
| Precision measuring matrix control sample | Repeated measures of a positive human C9orf72 CSF sample should have CV ≤ 20%. | Figure 3G. Supplementary table 7. |
| Dilutional parallelism | At least three of diluted samples within the assay's range should have DFT within ± 30.0% | Figure 3H. Supplementary table 8 and supplementary figure 1. |
| Freeze-thaw stability | Freeze thaw stability of matrix control QC. CV ≤ 25% and DFT $\leq \pm$ 30%.<br>Freeze thaw stability of calibrators CV ≤ 20%. | Figure 3G. Supplementary table 9 and supplementary table 10. |
| Haemoglobin tolerance | Assay should tolerate low levels of Haemoglobin within ± 20%. | Figure 3I and 3J. Supplementary figure 2. |

**Figure 3.**
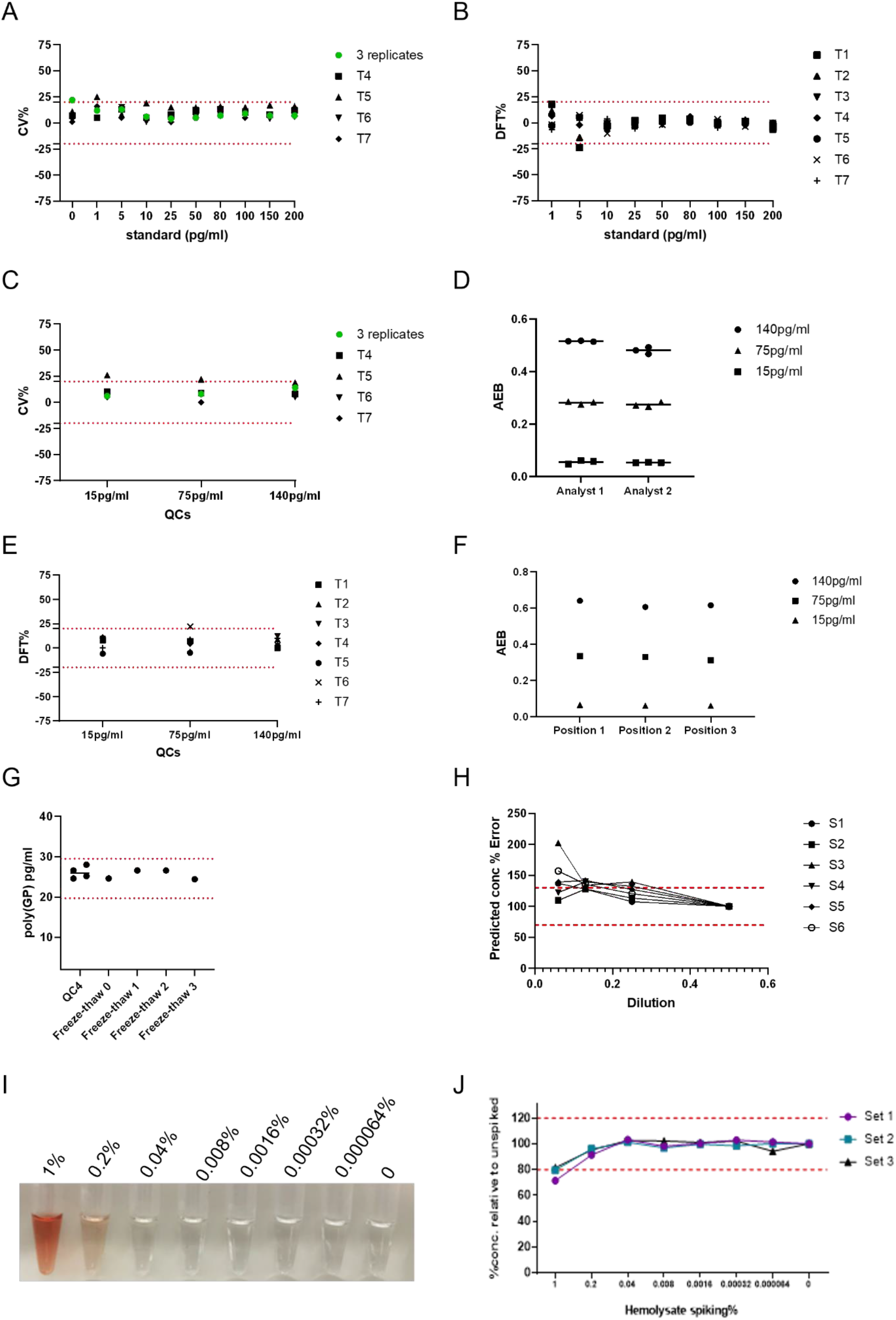
CSF poly(GP) Simoaassay qualification. 10-point standard curves ranging from 200 to 1 pg/ml and 3 quality control (QC) samples (15 pg/ml, 75 pg/ml 140 pg/ml) were prepared using GST-GP32 peptide and measured on 7 independent assays. **A)** The coefficient variation (CV) was measured for each standard, calculating first the CV for 3 initial assays (green dot) and then comparing subsequent assays to the average signal from those 3 assays. Red dotted line at +/- 20% acceptance level. **B)** The difference from total (DFT) calculated for each standard across 7 independent assays. DFT= % difference between predicted concentration and actual concentration of calibrators. Red dotted lines at +/- 20% acceptance level. **C)** CVs for QC samples across 7 independent assays. Green dot displaying the CV from the 3 initial assays. Red dotted lines at +/- 20% acceptance level. **D)** The Simoa assay signal, average number of enzyme labels per bead (AEB), measured for QCs prepared by 2 different analysts. Each analyst prepared 3 independent sets of QCs. **E)** DFTs calculated for QC samples run in 7 independent assays. Red dotted lines at +/- 20% acceptance level. **F)** Intra-plate variability assessed by measuring QCs in 3 different positions across a single assay plate. **G)** Human *C9orf72* CSF donor sample (QC4) measured in 4 independent assays, showing high precision. Furthermore, QC4 underwent 0, 1, 2, or 3 freeze-thaw cycles prior to measure in a single assay. Red dotted lines at +/- 20% acceptance level from the fresh measured QC4 sample. **H)** Dilutional parallelism measured using 6 *C9orf72* CSF samples serially diluted, using 1 in 2 dilution as anchor. Predicted concentration % error was calculated comparing the adjusted predicted concentration at each dilution to the concentration of the 1 in 2 diluted sample (set to 100%). Red dotted lines denote +/- 30% from the expected predicted concentration. **I)** Photo of CSF spiked with hemolysate ranging from 1% to 0.000064%. **J)** CSF was spiked with hemolysate and serially diluted to give range of equivalent % hemolysate. CSF was also spiked with 50 pg/ml GST-GP32 and poly(GP) concentration measured using Simoa assay. Three sets were assayed and % error in predicted concentration was plotted for each sample. Red dotted lines at +/- 20% from expected poly(GP) concentration.

Dilutional parallelism was assessed by running CSF from six *C9orf72* expansion positive donors either neat, 1:2, 1:4, 1:8 and 1:16 in diluent A. Poly(GP) was detected above background in all dilutions.

Using 1:2 as an anchor point the average % error of 4 out of 6 samples had <30% error at 1:4 dilution, passing qualification criteria (Figure 3H). The percentage error increased above 30% for the majority of samples at 1:8 and 1:16 (Table S8 and Figure S2). We chose to run samples at 1:2 dilution and recommend further assessment of parallelism within trials with more samples. Freeze-thaw stability of poly(GP) in CSF was tested using QC4 and measuring poly(GP) after 1, 2, and 3 freeze-thaw cycles. The signal and concentration measured had CV s of 4% and 5% respectively indicating no effect of freeze-thaw on detection of endogenous poly(GP) (Figure 3G and Table S9). The freeze thaw stability of the standard (GST-GP32) was also assessed after 1, 2, or 3 freeze thaw cycles. Eight of the calibrators passed criteria with CV <20% and DFT <20% (Table S10). The lowest standard curve point, 1 pg/ml gave a higher DFT after 3 freeze thaw cycles, but this is explained by the higher CV in signal measured for the blank in this set of calibrators, and we therefore concluded that it is unlikely that up to three freeze-thaw cycles affects the signal from GST-GP32.

During CSF collection it is possible for blood to contaminate the collected CSF. We tested if haemoglobin interfered with poly(GP) detection. We spiked a range of hemolysate concentrations (Figure 3I) into control CSF and spiked with either 5 pg/ml or 50 pg/ml GST-GP32. 5 pg/ml GST-GP32 spiked in CSF was not affected by any of the hemolysate concentrations tested (Figure S3). The measurement of 50 pg/ml GST-GP32 spiked in CSF was inhibited (>20%) by addition of 1% hemolysate (Figure 3J). At this concentration of haemoglobin, the CSF is visibly red (Figure 3I), so samples can be excluded from analysis by appearance if required. Note, none of the CSF samples measured in this study had a red or pink appearance.

### Measurement of poly(GP) in CSF from *C9orf72* expansion carriers using the optimised, qualified Simoa assay

We used this sensitive, qualified assay to measure poly(GP) in a cohort of CSF from healthy controls (N=15) and *C9orf72* expansion positive donors (N=40). The assay signal from the lowest *C9orf72* case had signal/noise 8-fold over the average signal from control samples, showing a clear separation from signals of control CSF (Figure 4A). On average the signal to noise of *C9orf72* cases versus controls was 38-fold. Poly(GP) in CSF from healthy donors was below detection level for 13 out of 15 samples or below LLOQ of the assay for the remaining 2 out of 15 cases. As poly(GP) was detected above LLOQ in all *C9orf72* cases and in no healthy controls, sensitivity and specificity were both 100%. Poly(GP) measures ranged from 6 – 148 pg/ml in *C9orf72* expansion positive donors. Despite the increased sensitivity of this Simoa assay, the levels of poly(GP) were not statistically different between presymptomatic and symptomatic *C9orf72* expansion positive donors (p=0.1348 Mann-Whitney test), although we observed the same trend observed by others towards higher levels in symptomatic cases^15,22,33^ (Figure 4B). We found no difference in poly(GP) levels between male and female *C9orf72* expansion positive donors (Figure S4A). We found no correlation between CSF poly(GP) levels and age of onset of symptomatic *C9orf72* expansion positive donors (N=15) (Figure 4C). Interestingly there was a significant, moderate positive correlation (r= 0.3643) between age at donation and poly(GP) measured in CSF, analysing all 40 *C9orf72* expansion positive cases (Figure 4D). However, if the case with the highest poly(GP) level is removed from analysis the P value changes to P=0.0522.

**Figure 4.**
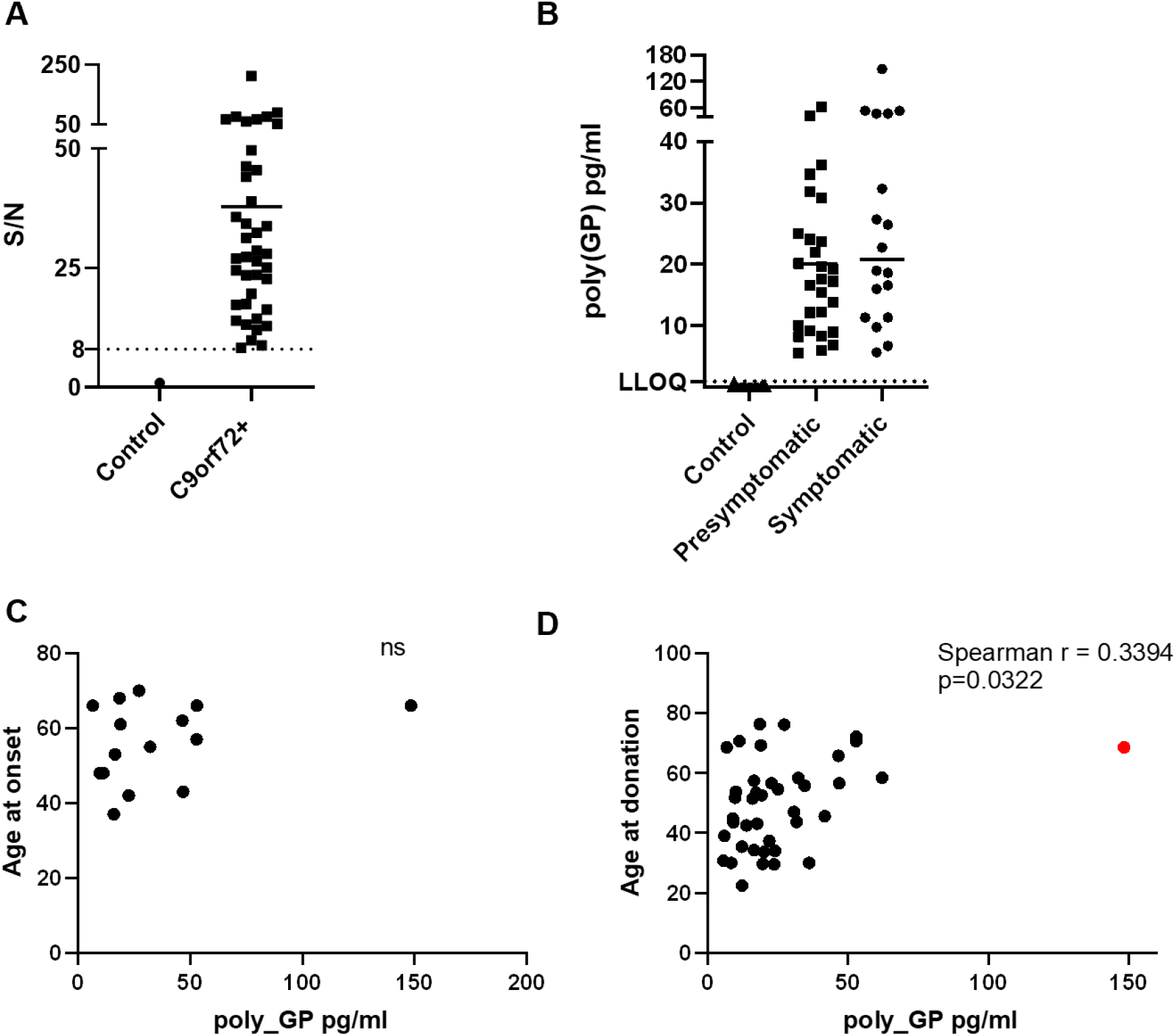
Poly(GP) levels in CSF from C9orf72 expansion carriers. Poly(GP) levels in CSF from 25 presymptomatic *C9orf72* expansion carriers, 15 symptomatic *C9orf72* carriers and 15 healthy aged matched controls were measured using our optimised Simoa HD-X assay. **A)** Signal/noise (S/N) was calculated by dividing the mean AEB signal from duplicate measures of 40 *C9orf72* expansion carriers, by the mean AEB signal of CSF from all 15 healthy controls (plotted here as 1). *C9orf72* expansion carriers had poly(GP) assay signals distinct from healthy controls, with all S/N values above 8. **B)** Comparison of poly(GP) levels in presymptomatic and symptomatic *C9orf72* expansion carriers. Each data point is the average from a duplicate measure from each donor, with bar at mean for each group. Lower Limit of Quantification (LLOQ) at 1 pg/ml is shown with dotted line. There is no statistical difference in poly(GP) levels between presymptomatic and symptomatic *C9orf72* expansion carriers (Mann–Whitney U test). **C)** Age of onset plotted against poly(GP) pg/ml in CSF for 15 symptomatic *C9orf72* expansion carriers. ns = not significant, no correlation found (Spearman r). **D)**Age at donation plotted against CSF poly(GP) levels. Red dot indicates high poly(GP) CSF case, which if removed increases p value to p=0.0522.

Where data was available we also tested for correlations between CSF poly(GP) levels and both total brain and lobar volumes. No correlation was found, analysing all *C9orf72* expansion carriers or selecting symptomatic cases only (Figure S5), consistent with a previous report^33^. Plasma neurofilament light chain (NfL) is a known biomarker of neurodegeneration. Plasma levels of NfL were measured in 18 of the *C9orf72* expansion carrier CSF donors (including 8 symptomatic donors). As expected, plasma NfL levels were significantly higher in symptomatic carriers (Figure S6A). No correlation was found between CSF poly(GP) and plasma NfL levels analysing the small sample of 8 symptomatic cases (Figure S6B).

We next optimised our poly(GP) Simoa assay for analysis of plasma. Despite the high sensitivity of the Simoa platform we were unable to detect poly(GP) in plasma from *C9orf72* expansion positive donors. Signals were below LLOQ and there was no difference between control- and *C9orf72*-positive signals (Figure S6C). The two cases of plasma from *C9orf72* expansion carriers which had higher AEB signals were not the same donors with higher than average CSF poly(GP), and there was no correlation between plasma AEB signal and poly(GP) measured in matched CSF samples (Figure S6D). There is a predicted 200-fold drop in concentration of NfL measured between CSF and plasma. The levels of poly(GP) in CSF were on average 26 pg/ml, so if a similar reduction is observed for poly(GP) a platform capable of detecting in femtogram range maybe required to measure poly(GP) in plasma.

## Discussion

We describe the development and qualification of a sensitive Simoa assay for poly(GP) DPRs in CSF. Multiple antibodies were assessed and compared in combinations in a Homebrew Simoa assay, identifying differences in performance across antibody combinations. In our experience not all polyclonal antibodies behave the same, even when the same peptide sequence was used for antigen. We tested the performance of a monoclonal antibody as both capture and detector in a Homebrew Simoa assay. Unfortunately, the monoclonal antibody tested here, did not perform as well as a detector antibody as the polyclonal antibodies, with much higher predicted LLOQs. The reason for this difference is unclear, but the different polyclonal antibodies may recognise different secondary structures of poly(GP). As part of assay development we transferred our Home brew poly(GP) Simoa assay from HD-1 analyser to the newer HD-X model. We found the assay required re-optimisation, with a change in sample diluent affecting assay performance the most.

After choosing antibodies as capture and detector the Simoa assay was optimised, testing number of washes, concentration of detector antibody, enzyme concentration, and addition of helper-beads.

After selecting the best performing conditions, the assay was qualified to determine its suitability for use in a clinical trial setting. Standard curves ranging from 200 pg/ml to 1 pg/ml were made using GST-GP32 peptide and run in 7 independent assays, by two different analysts. Accuracy and precision passed criteria with DFT and CVs below 20% for all standard curve points. The assay was found to be precise when testing QC samples both inter-plate and intra-plate. Preparing this assay for use in clinical trials, we tested dilutional parallelism, using 6 CSF samples from *C9orf72* expansion carriers. The matrix effect of CSF was evident and we would recommend minimum required dilution of clinical samples 1:2, with ability to dilute up to 1:8 before parallelism is lost and % error exceeds +/-30%. Further assessment of parallelism should take place in trial assessing more samples. In case samples are re-assessed we measured the freeze-thaw stability of a *C9orf72* positive CSF sample.

Precision and accuracy were maintained across 3 freeze-thaw cycles with DFT and CVs below 10%, indicating clinical samples can withstand freeze-thaw. During CSF collection it is possible that some patient blood can contaminate the CSF. Here, we assessed if haemoglobin can interfere with detection of poly(GP). Our assay was not affected by haemoglobin interference, with only the most severely contaminated CSF sample (+1% hemolysate) showing altered poly(GP) detection. This level of CSF blood contamination is visible by eye and can be removed from analysis.

We used our qualified poly(GP) assay to analyse CSF from a small cohort of CSF samples provided by GENFI, including 15 healthy controls and 40 *C9orf72* expansion carriers. Similar to previously published studies^15,22,33^, our assay was able to distinguish controls and *C9orf72* expansion carriers. In this cohort we had 100% sensitivity and 100% specificity with poly(GP) measured in CSF from all *C9orf72* expansion carriers, whilst controls either measured below detection (13/15) or below limit of quantification (2/15), determined at 1 pg/ml. *C9orf72* expansion carriers had a range of poly(GP) from 6 -148 pg/ml, with all positive sample signals at least 8-fold higher than control signals, showing a clear separation of controls from *C9orf72* expansion samples. Previous studies using MSD immunoassays reported the average CSF polyGP signal to be in the low nanogram range ^15,33^, while our assay gives average polyGP levels in the low-medium picogram range. This difference may be attributed to the different calibrators used in the studies, as we have noted that the same antibody can report different concentrations depending on the calibrator used. In our cohort of samples we found, similar to previous studies, that compared to presymptomatic carriers, symptomatic carriers had higher levels of poly(GP) comparing median levels, but this difference was not significant ^15,22^.

Meeter *et al*. (2018) found levels in symptomatic carriers were significantly higher^33^. This may be due to the larger cohort size tested with more symptomatic donors with higher than average poly(GP) levels included. Within our small cohort there was one symptomatic *C9orf72* carrier with much higher poly(GP) levels than the rest. Age at onset (66 years) and age at donation (68 years) were both within 1 standard deviation from the mean of other symptomatic donors, indicating no effect of higher levels of poly(GP) on these parameters. We did not have repeat length data for this cohort, although given the variability in repeat length between different tissues in the body it would be difficult to interpret repeat length data determined from blood DNA. Lehmer *et al*. (2017) found no correlation between repeat size and CSF poly(GP) levels in 11 cases where DNA was available ^22^.

Should post-mortem tissue become available from donors in this cohort, it would be interesting to determine repeat length from brain tissue as well as measure propensity of DPR aggregates in the brain to see if poly(GP) CSF levels reflected aggregate burden.

Similar to previous studies we found no correlations between CSF poly(GP) levels and clinical features including; gender, age of onset or brain volume, analysing either total *C9orf72* cases or just symptomatic *C9orf72* carriers^15,22,33^. We did observe a correlation between CSF poly(GP) levels and age at donation, which is potentially consistent with a relationship between *C9orf72* expansion length and age at DNA sample collection^34^. We analysed NfL levels in a subset of donor matched plasma samples. As expected, symptomatic carriers had higher NfL plasma levels than presymptomatic or controls. As in previous studies that measured NfL in CSF^22,33^, NfL plasma levels did not correlate with poly(GP) CSF levels. Despite the ability of the Simoa assays to detect at single - molecule levels, we were unable to measure poly(GP) in donor matched plasma samples. Signals for all samples were below quantification and did not correlate with poly(GP) CSF levels. If poly(GP) produced in the brain is present in plasma it will require a more sensitive assay platform and a better understanding of potential matrix effects. Neurofilament assays have been shown to be able to utilise either blood or CSF to monitor levels of neurodegeneration^35–37^. In contrast, this qualified poly(GP) assay will specifically determine target engagement in an upcoming ASO clinical trial, which may occur prior to changes in levels of neurodegeneration. In summary, we show utility of the Simoa HD-X platform for detecting poly(GP) in the CSF of people with a *C9orf72* expansion, with assay reliability good enough to be used for target engagement analysis in clinical trials directly targeting *C9orf72* repeat containing transcripts.

## Data Availability

All data produced in the present study are available upon reasonable request to the authors.

## Acknowledgments

We thank the GENFI research participants for their contribution to the study. This work was funded by Wave Life Sciences, the European Research Council (ERC) under the European Union’s Horizon 2020 research and innovation programme (648716 - C9ND) (AMI), the UK Dementia Research Institute, which receives its funding from UK DRI Ltd, funded by the UK Medical Research Council, Alzheimer’s Society and Alzheimer’s Research UK. The Dementia Research Centre is supported by Alzheimer’s Research UK, Alzheimer’s Society, Brain Research UK, and The Wolfson Foundation. This work was supported by the NIHR UCL/H Biomedical Research Centre, the Leonard Wolfson Experimental Neurology Centre (LWENC) Clinical Research Facility and the NIHR Cambridge Biomedical Research Centre (BRC-1215-20014). The views expressed are those of the authors and not necessarily those of the NIHR or the Department of Health and Social Care. HZ is a Wallenberg Scholar. Simoa instruments used were funded by Wellcome Trust, Fidelity International Foundation and UK DRI. JDR is supported by the Miriam Marks Brain Research UK Senior Fellowship and has received funding from an MRC Clinician Scientist Fellowship (MR/M008525/1) and the NIHR Rare Disease Translational Research Collaboration (BRC149/NS/MH). This work was also supported by the MRC UK GENFI grant (MR/M023664/1), the Bluefield Project and the JPND GENFI-PROX grant (2019-02248). This work was also supported by the MRC UK GENFI grant (MR/M023664/1). Several authors of this publication are members of the European Reference Network for Rare Neurological Diseases - Project ID No 739510.

## APPENDIX A

***List of GENFI consortium authors:***

Sónia Afonso^1^, Maria Rosario Almeida^2^, Sarah Anderl-Straub^3^, Christin Andersson^4^, Anna Antonell^5^, Silvana Archetti^6^, Andrea Arighi^7^, Mircea Balasa^8^, Myriam Barandiaran^9^, Nuria Bargalló^10^, Robart Bartha^11^, Benjamin Bender^12^, Alberto Benussi^13^, Maxime Bertoux^14,15^, Anne Bertrand^16,17^, Valentina Bessi^19^, Sandra Black^20^, Martina Bocchetta^21^, Sergi Borrego-Ecija^8^, Jose Bras^22^, Alexis Brice^16^, Rose Bruffaerts^23,24^, Agnès Camuzat^8^, Marta Cañada^25^, Valentina Cantoni^13^, Paola Caroppo^26^, David Cash^13^, Miguel Castelo-Branco^2^, Olivier Colliot^16,17^, Rhian Convery^13^, Thomas Cope^27^, Adrian Danek^18^, Vincent Deramecourt^29,30,31^, Giuseppe Di Fede^26^, Alina Díez^31^, Diana Duro^2^, Chiara Fenoglio^7^, Camilla Ferrari^32^, Catarina B. Ferreira^33^, Nick Fox^21^, Morris Freedman^34^, Giorgio Fumagalli^7^, Aurélie Funkiewiez^16,35^, Alazne Gabilondo^31^, Roberto Gasparotti^37^, Serge Gauthier^38^, Stefano Gazzina^39^, Giorgio Giaccone^26^, Ana Gorostidi^36^, Lisa Graf^40^, Caroline Greaves^21^, Rita Guerreiro^22^, Tobias Hoegen^41^, Begoña Indakoetxea^9^, Vesna Jelic^42^, Lize Jiskoot^43^, Ron Keren^44^, Gregory Kuchcinski^27,28,29^, Tobias Langheinrich^45,46^, Isabelle Le Ber^16,35,47^, Thibaud Lebouvier^27,28,29^, Maria João Leitão^48^, Johannes Levin^,18,19,50^, Albert Lladó^8^, Gemma Lombardi^32^, Jolina Lombardi^3^, Sandra Loosli^51^, Carolina Maruta^52^, Simon Mead^53^, Gabriel Miltenberger^2^, Rick van Minkelen^54^, Sara Mitchell^20^, Fermin Moreno^9^, Benedetta Nacmias^32,55^, Annabel Nelson^21^, Jennifer Nicholas^56^, Linn Öijerstedt57,58, Janne M. Papma43, Florence Pasquier14,15, Georgia Peakman21, Yolande Pijnenburg59, Cristina Polito60, Enrico Premi^61^, Sara Prioni^26^, Catharina Prix^51^, Veronica Redaelli^26^, Daisy Rinaldi^16,35,47^, Tim Rittman^27^, Ekaterina Rogaeva^62^, Pedro Rosa-Neto^63^, Giacomina Rossi^26^, Martin Rossor^21^, Isabel Santana^64^, Beatriz Santiago^26^, Dario Saracino^16,35,47^, Elio Scarpini^7^, Sonja Schönecker^51^, Rachelle Shafei^21^, Christen Shoesmith^65^, Sandro Sorbi^32,55^, Miguel Tábuas-Pereira^66^, Fabrizio Tagliavini^26^, Mikel Tainta^31^, Ricardo Taipa^67^, David Tang-Wai^68^, David L Thomas^69^, Paul Thompson^70^, Carolyn Timberlake^27^, Pietro Tiraboschi^26^, Emily Todd^21^, Philip Van Damme^71^, Mathieu Vandenbulcke^72^, Ana Verdelho^73^, Jorge Villanua^74^, Jason Warren^21^, Carlo Wilke^40^, Elisabeth Wlasich^41^, Miren Zulaica^36^

## List of GENFI consortium affiliations

1. Instituto Ciencias Nucleares Aplicadas a Saude, Universidade de Coimbra, Coimbra, Portugal.
2. Faculty of Medicine, University of Coimbra, Coimbra, Portugal.
3. Department of Neurology, University of Ulm, Ulm, Germany.
4. Department of Clinical Neuroscience, Karolinska Institutet, Stockholm, Sweden.
5. Alzheimer’s disease and Other Cognitive Disorders Unit, Neurology Service, Hospital Clínic, Barcelona, Spain.
6. Biotechnology Laboratory, Department of Diagnostics, ASST Brescia Hospital, Brescia, Italy.
7. Fondazione IRCCS Ca’ Granda Ospedale Maggiore Policlinico, Neurodegenerative Diseases Unit, Milan, Italy; University of Milan, Centro Dino Ferrari, Milan, Italy.
8. Alzheimer’s disease and Other Cognitive Disorders Unit, Neurology Service, Hospital Clínic, Barcelona, Spain.
9. Cognitive Disorders Unit, Department of Neurology, Donostia University Hospital, San Sebastian, Gipuzkoa, Spain; Neuroscience Area, Biodonostia Health Research Insitute, San Sebastian, Gipuzkoa, Spain.
10. Imaging Diagnostic Center, Hospital Clínic, Barcelona, Spain.
11. Department of Medical Biophysics, The University of Western Ontario, London, Ontario, Canada; Centre for Functional and Metabolic Mapping, Robarts Research Institute, The University of Western Ontario, London, Ontario, Canada.
12. Department of Diagnostic and Interventional Neuroradiology, University of Tübingen, Tübingen, Germany.
13. Centre for Neurodegenerative Disorders, Department of Clinical and Experimental Sciences, University of Brescia, Italy.
14. Inserm 1172, Lille, France.
15. CHU, CNR-MAJ, Labex Distalz, LiCEND Lille, France.
16. Sorbonne Université, Paris Brain Institute – Institut du Cerveau – ICM, Inserm U1127, CNRS UMR 7225, AP-HP - Hôpital Pitié-Salpêtrière, Paris, France.
17. Centre pour l’Acquisition et le Traitement des Images, Institut du Cerveau et la Moelle, Paris, France.
18. Department of Neurology, Ludwig-Maximilians Universität München, Munich, Germany.
19. Department of Neuroscience, Psychology, Drug Research and Child Health, University of Florence, Florence, Italy.
20. Sunnybrook Health Sciences Centre, Sunnybrook Research Institute, University of Toronto, Toronto, Canada.
21. Dementia Research Centre, Department of Neurodegenerative Disease, UCL Institute of Neurology, Queen Square, London, UK.
22. Center for Neurodegenerative Science, Van Andel Institute, Grand Rapids, Michigan, MI 49503, USA.
23. Laboratory for Cognitive Neurology, Department of Neurosciences, KU Leuven, Leuven, Belgium.
24. Biomedical Research Institute, Hasselt University, 3500 Hasselt, Belgium.
25. CITA Alzheimer, San Sebastian, Gipuzkoa, Spain.
26. Fondazione IRCCS Istituto Neurologico Carlo Besta, Milano, Italy.
27. Department of Clinical Neuroscience, University of Cambridge, Cambridge, UK.
28. Univ Lille, France.
29. Inserm 1172, Lille, France.
30. CHU, CNR-MAJ, Labex Distalz, LiCEND Lille, France.
31. Neuroscience Area, Biodonostia Health Research Insitute, San Sebastian, Gipuzkoa, Spain.
32. Department of Neuroscience, Psychology, Drug Research and Child Health, University of Florence, Florence, Italy.
33. Laboratory of Neurosciences, Institute of Molecular Medicine, Faculty of Medicine, University of Lisbon, Lisbon, Portugal.
34. Baycrest Health Sciences, Rotman Research Institute, University of Toronto, Toronto, Canada
35. Centre de référence des démences rares ou précoces, IM2A, Département de Neurologie, AP-HP - Hôpital Pitié-Salpêtrière, Paris, France
36. Neuroscience Area, Biodonostia Health Research Insitute, San Sebastian, Gipuzkoa, Spain.
37. Neuroradiology Unit, University of Brescia, Brescia, Italy.
38. Alzheimer Disease Research Unit, McGill Centre for Studies in Aging, Department of Neurology & Neurosurgery, McGill University, Montreal, Québec, Canada.
39. Neurology, ASST Brescia Hospital, Brescia, Italy.
40. Department of Neurodegenerative Diseases, Hertie-Institute for Clinical Brain Research and Center of Neurology, University of Tübingen, Tübingen, Germany.
41. Neurologische Klinik, Ludwig-Maximilians-Universität München, Munich, Germany.
42. Division of Clinical Geriatrics, Karolinska Institutet, Stockholm, Sweden.
43. Department of Neurology, Erasmus Medical Center, Rotterdam, Netherlands.
44. The University Health Network, Toronto Rehabilitation Institute, Toronto, Canada.
45. Division of Neuroscience and Experimental Psychology, Wolfson Molecular Imaging Centre, University of Manchester, Manchester, UK.
46. Manchester Centre for Clinical Neurosciences, Department of Neurology, Salford Royal NHS Foundation Trust, Manchester, UK.
47. Département de Neurologie, AP-HP - Hôpital Pitié-Salpêtrière, Paris, France
48. Centre of Neurosciences and Cell Biology, Universidade de Coimbra, Coimbra, Portugal.
49. German Center for Neurodegenerative Diseases (DZNE), Munich, Germany.
50. Munich Cluster of Systems Neurology (SyNergy), Munich, Germany.
51. Neurologische Klinik, Ludwig-Maximilians-Universität München, Munich, Germany.
52. Laboratory of Language Research, Centro de Estudos Egas Moniz, Faculty of Medicine, University of Lisbon, Lisbon, Portugal.
53. MRC Prion Unit, Department of Neurodegenerative Disease, UCL Institute of Neurology, Queen Square, London, UK.
54. Department of Clinical Genetics, Erasmus Medical Center, Rotterdam, Netherlands.
55. IRCCS Fondazione Don Carlo Gnocchi, Florence, Italy.
56. Department of Medical Statistics, London School of Hygiene and Tropical Medicine, London, UK.
57. Center for Alzheimer Research, Division of Neurogeriatrics, Department of Neurobiology, Care Sciences and Society, Bioclinicum, Karolinska Institutet, Solna, Sweden.
58. Unit for Hereditary Dementias, Theme Aging, Karolinska University Hospital, Solna, Sweden.
59. Amsterdam University Medical Centre, Amsterdam VUmc, Amsterdam, Netherlands.
60. Department of Biomedical, Experimental and Clinical Sciences “Mario Serio”, Nuclear Medicine Unit, University of Florence, Florence, Italy.
61. Stroke Unit, ASST Brescia Hospital, Brescia, Italy.
62. Tanz Centre for Research in Neurodegenerative Diseases, University of Toronto, Toronto, Canada
63. Translational Neuroimaging Laboratory, McGill Centre for Studies in Aging, McGill University, Montreal, Québec, Canada.
64. University Hospital of Coimbra (HUC), Neurology Service, Faculty of Medicine, University of Coimbra, Coimbra, Portugal; Center for Neuroscience and Cell Biology, Faculty of Medicine, University of Coimbra, Coimbra, Portugal.
65. Department of Clinical Neurological Sciences, University of Western Ontario, London, Ontario, Canada.
66. Neurology Department, Centro Hospitalar e Universitario de Coimbra, Coimbra, Portugal.
67. Neuropathology Unit and Department of Neurology, Centro Hospitalar do Porto - Hospital de Santo António, Oporto, Portugal.
68. The University Health Network, Krembil Research Institute, Toronto, Canada.
69. Neuroimaging Analysis Centre, Department of Brain Repair and Rehabilitation, UCL Institute of Neurology, Queen Square, London, UK.
70. Division of Neuroscience and Experimental Psychology, Wolfson Molecular Imaging Centre, University of Manchester, Manchester, UK.
71. Neurology Service, University Hospitals Leuven, Belgium; Laboratory for Neurobiology, VIB-KU Leuven Centre for Brain Research, Leuven, Belgium.
72. Geriatric Psychiatry Service, University Hospitals Leuven, Belgium; Neuropsychiatry, Department of Neurosciences, KU Leuven, Leuven, Belgium.
73. Department of Neurosciences and Mental Health, Centro Hospitalar Lisboa Norte - Hospital de Santa Maria & Faculty of Medicine, University of Lisbon, Lisbon, Portugal.
74. OSATEK, University of Donostia, San Sebastian, Gipuzkoa, Spain.

