## Supplemental materials for "Development of a sensitive trial-ready poly(GP) CSF biomarker assay for *C9orf72*-associated frontotemporal dementia and amyotrophic lateral sclerosis"

**Supplemental Information**

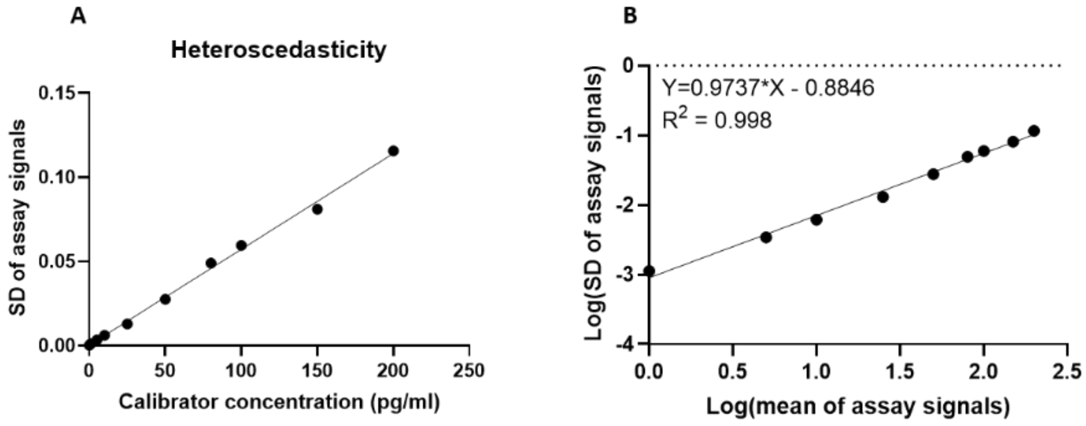


C

| 4PL- 1/y^2 | | 5PL- 1/y^2 | | 4PL- 1/y^1.9474 | | 5PL- 1/y^1.9474 | | 4PL- 1/y^none | | 5PL- 1/y^none | |
|---|---|---|---|---|---|---|---|---|---|---|---|
| **%RE** | **CV%** | %RE | CV% | %RE | CV% | %RE | CV% | %RE | CV% | %RE | CV% |
| **-2.21** | **2.2** | -2.21 | 2.1 | -0.50 | 0.8 | -0.79 | 1.0 | -0.50 | 0.7 | -0.71 | 1.1 |
| **0.19** | **2.1** | 0.19 | 2.1 | 1.14 | 2.0 | 1.81 | 2.3 | 13.62 | 39.4 | 1.81 | 2.3 |
| **-0.59** | **2.5** | -0.59 | 2.5 | -0.71 | 2.7 | 0.76 | 2.5 | -0.71 | 2.8 | 0.56 | 2.6 |
| **3.32** | **2.1** | 3.34 | 2.1 | 2.59 | 1.9 | 3.86 | 2.6 | 2.50 | 1.9 | 3.79 | 2.7 |
| **1.37** | **2.3** | 1.49 | 2.4 | -0.77 | 2.7 | 0.17 | 4.9 | -1.03 | 2.3 | 0.20 | 4.9 |
| **-1.37** | **2.3** | 4.40 | 16.1 | -5.43 | 5.5 | -5.49 | 7.9 | -6.00 | 4.8 | -5.20 | 7.7 |
| **-2.89** | **4.5** | -2.81 | 4.6 | -7.09 | 9.4 | -7.93 | 12.0 | -7.86 | 9.1 | -7.36 | 11.7 |
| **-2.89** | **11.4** | -2.89 | 11.4 | -0.20 | 7.7 | 0.89 | 5.1 | -0.74 | 7.9 | 0.86 | 5.0 |
| **2.93** | **9.3** | 2.81 | 9.1 | *38.15* | *29.1* | *50.25* | *93.9* | *37.07* | *35.2* | *50.25* | *93.9* |

**Supplementary figure 1. Assessment of curve fitting.** A) Assessment of heteroscedasticity of data carried out by plotting standard deviation of assay signals (AEB) from the calibrator curve standards from 7 independent assays, against the calibrator concentration (pg/ml). B) To calculate weighting, linear regression was applied after plotting Log(standard deviation of assay signals) against Log(mean of assay signals) and the slope of the line (k) used in the formula: weighting= $1/Y^{2k}$. C) Curves were recalculated using 4PL and 5PL, with no weighting, 1.9474, or 2 weighting. Curve fits were assessed using criteria that % cumulative relative errors (RE%) and CV% for calibrators were +/- 15%, and RE% and CV% for anchor points (1 pg/ml) were +/- 20%. When 4PL $1/Y^2$ was used for curve fitting, all calibrator points passed these criteria and 4PL $1/Y^2$ was therefore chosen.

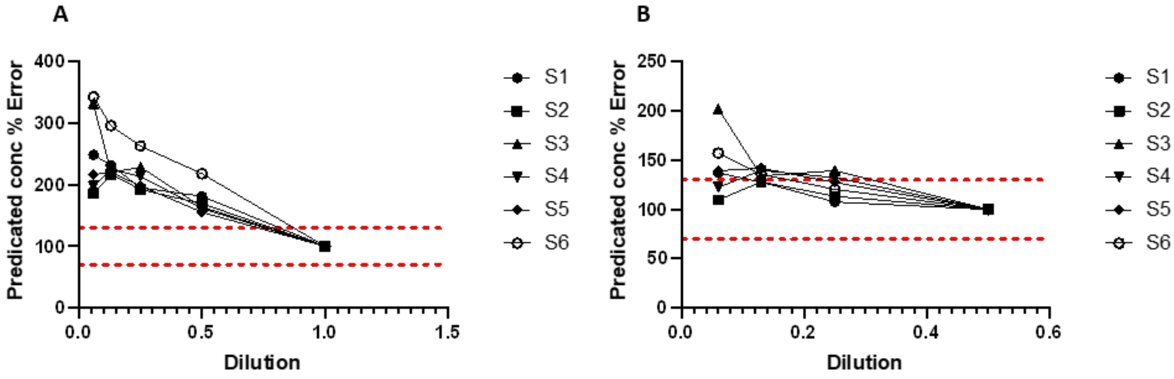


**Supplementary figure 2. Dilutional parallelism.** CSF from six *C9orf72* expansion positive donors was measured either neat, 1:2, 1:4, 1:8 and 1:16 diluted in diluent A. The mean AEB from duplicate measures was used to predict concentration at each dilution. **A**) The neat sample concentration was used as anchor and the % error was calculated comparing the adjusted predicted concentration at each dilution to the concentration of the neat sample. **B**) The 1:2 diluted sample used as anchor instead. Red dotted lines denote +/- 30% from the expected predicted concentration.

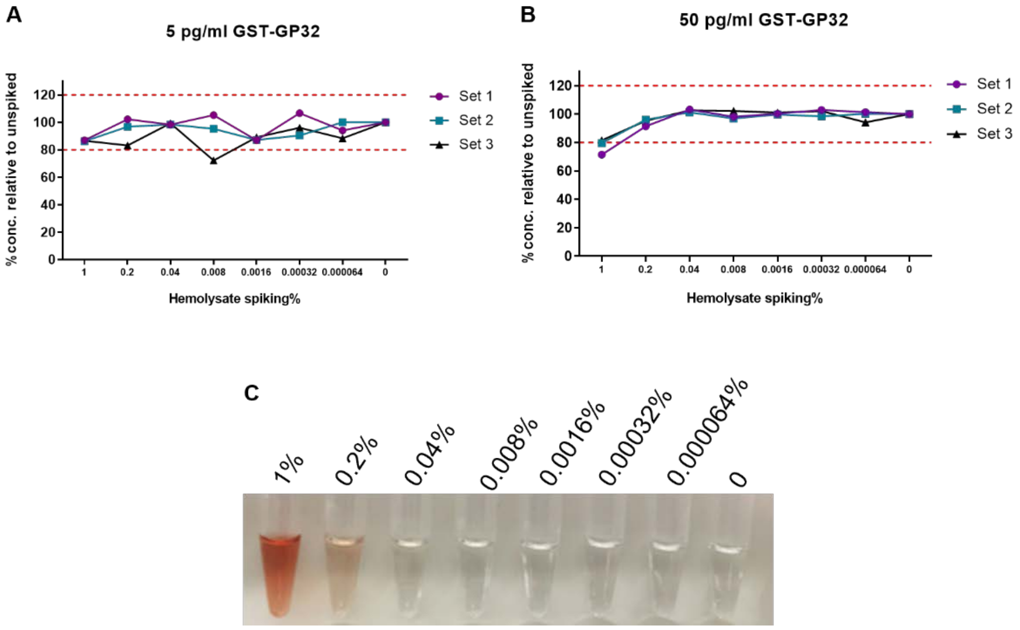


**Supplementary figure 3. Haemoglobin interference.** Control CSF was spiked with haemolysate and serial diluted to give range of equivalent % haemolysate. CSF was also spiked with either 5 pg/ml (A) or 50 pg/ml GST-GP32 (B) and poly(GP) concentration measured using Simoa assay. Three sets at each GST-GP32 concentration were assayed and % error in predicated concentration was plotted for each sample. Red dotted lines at +/- 20% from expected poly(GP) concentration. C) Visual appearance of CSF after haemolysate spiking.

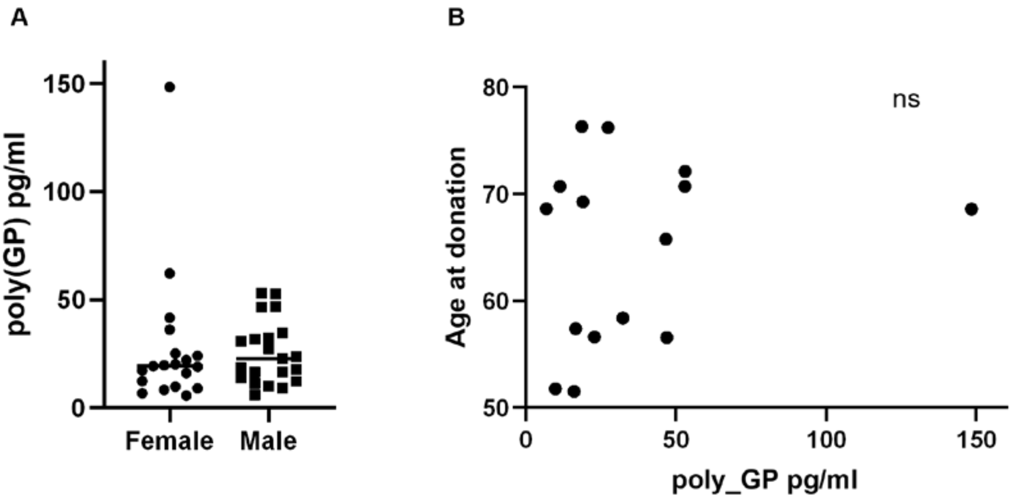


**Supplementary Figure 4. Analysis of poly(GP) CSF levels with clinical features. A)** No difference between female and male *C9orf72* expansion carriers in CSF poly(GP) levels. **B).** Age at visit for symptomatic *C9orf72* expansion carriers plotted against CSF poly(GP) levels.

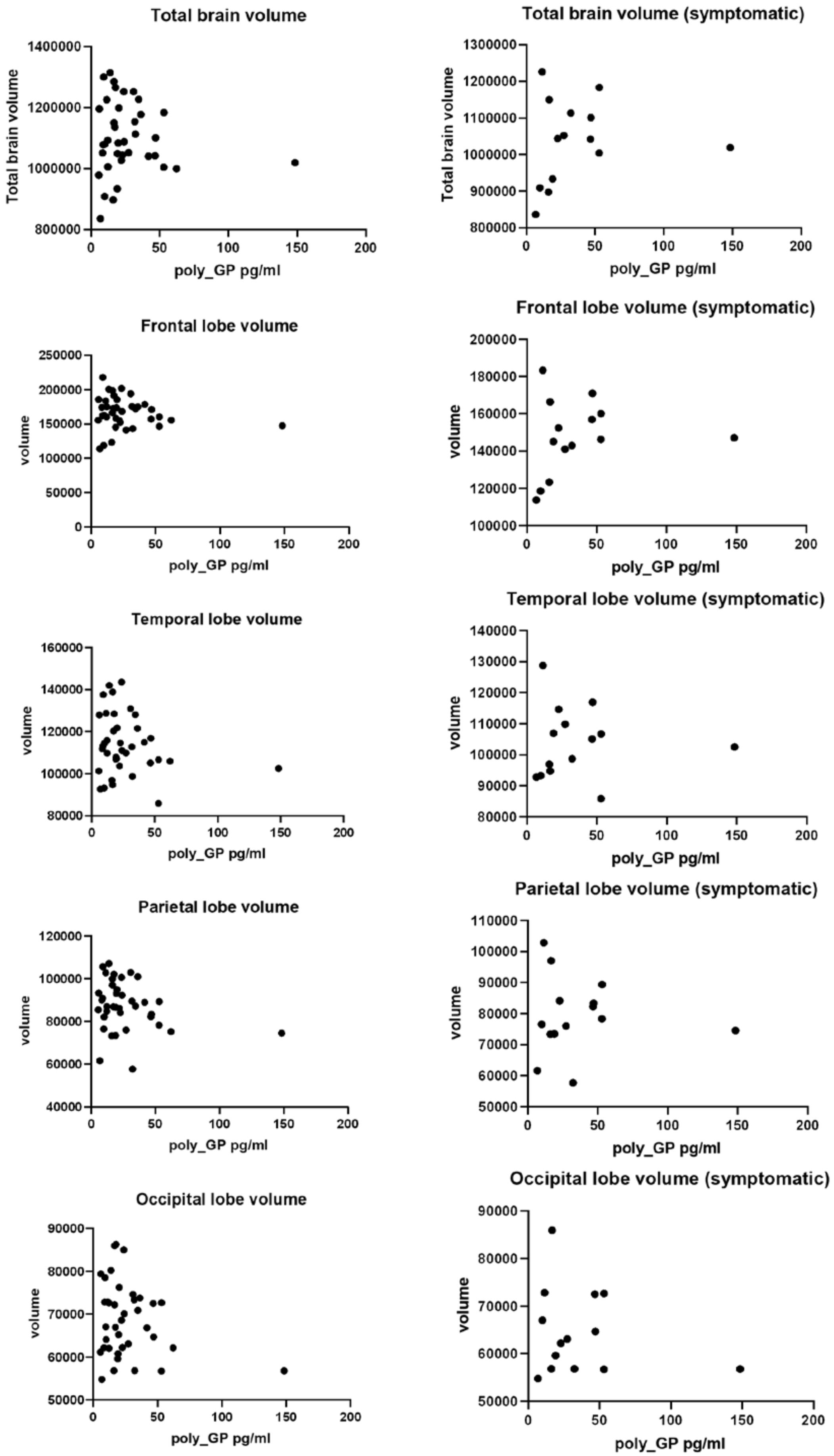
Total brain volume
Total brain volume
poly_GP pg/ml
Total brain volume (symptomatic)
Total brain volume
poly_GP pg/ml
Frontal lobe volume
volume
poly_GP pg/ml
Frontal lobe volume (symptomatic)
volume
poly_GP pg/ml
Temporal lobe volume
volume
poly_GP pg/ml
Temporal lobe volume (symptomatic)
volume
poly_GP pg/ml
Parietal lobe volume
volume
poly_GP pg/ml
Parietal lobe volume (symptomatic)
volume
poly_GP pg/ml
Occipital lobe volume
volume
poly_GP pg/ml
Occipital lobe volume (symptomatic)
volume
poly_GP pg/ml

**Supplementary Figure 5. Analysis of brain volume with poly(GP) CSF levels.** Left-hand side; total brain volume, temporal lobe, parietal lobe, occipital lobe and frontal lobe volumes against poly(GP) CSF levels from *C9orf72* expansion carriers (N=38). Right-hand side analysis of same regions from symptomatic *C9orf72* expansion carriers (N=14).

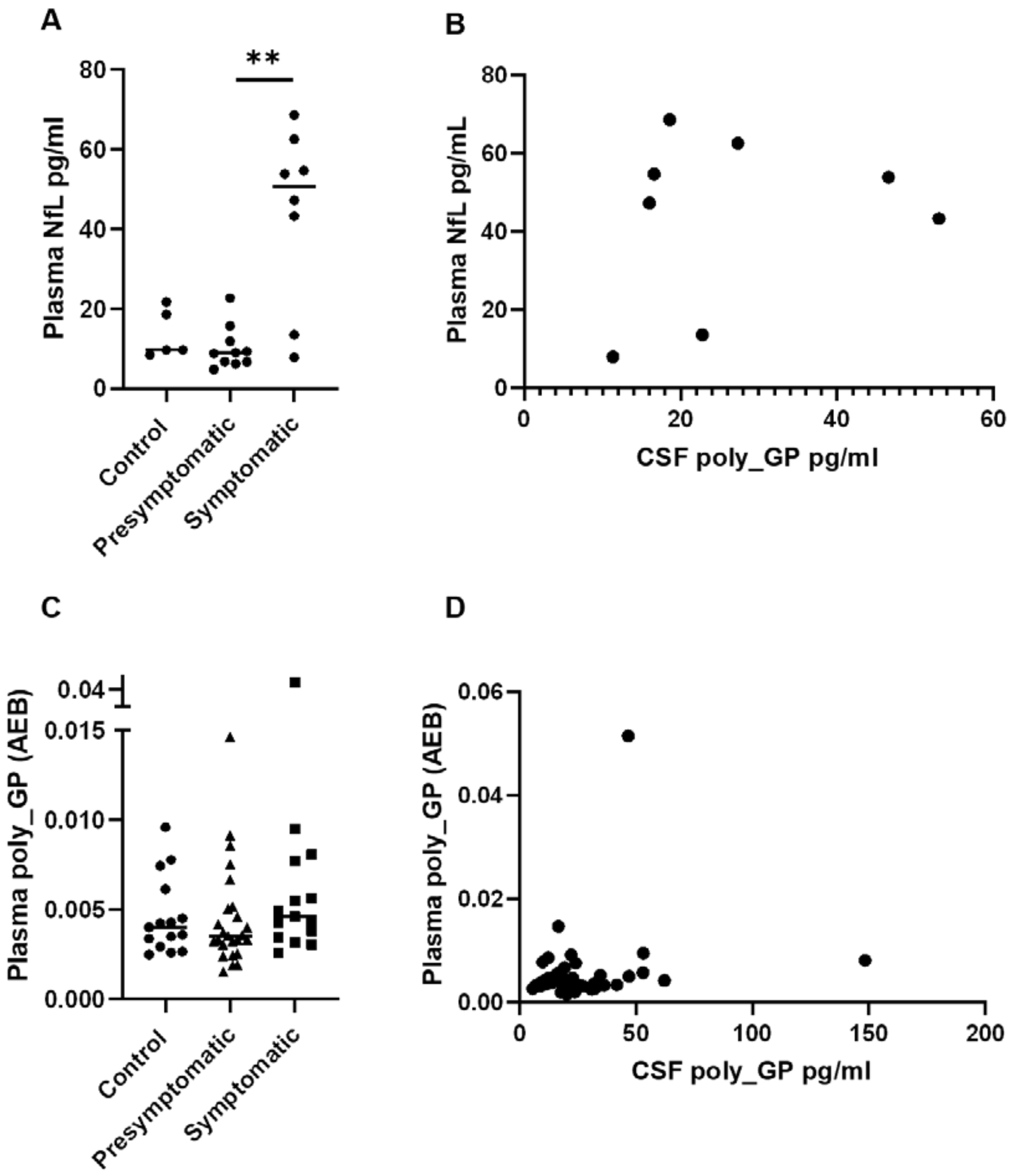


**Supplementary Figure 6. Analysis of plasma biomarkers from matched CSF donors.** Plasma samples from 5 controls, 10 presymptomatic and 8 symptomatic *C9orf72* expansion carriers whom also had poly(GP) CSF measured. **A)** Plasma NfL levels were significantly higher in symptomatic carriers compared to presymtomatic carriers (Kruskal Wallis and Dunn's multiple comparisons, ** p<0.01). **B)** No correlation was observed between plasma NfL levels and CSF poly(GP) levels in the available matched samples from 8 symptomatic cases. **C)** Raw AEB signals from Simoa assay optimised to measure poly(GP) in plasma. No difference was observed between controls or *C9orf72* expansion carriers. **D)** Raw AEB signals from plasma samples plotted against matched samples CSF poly(GP) levels.

| | Avg. from 3 independent assays | | Comp. TEST4 to average | | Comp. TEST5 to average | | Comp. TEST6 to average | | Comp. TEST7 to average | |
|---|---|---|---|---|---|---|---|---|---|---|
| Calibration curve | Analyst 1 | | Analyst 2 | | Analyst 2 | | Analyst 1 | | Analyst 2 | |
| pg/ml | Mean AEB | CV% | AEB | CV% comp. | AEB | CV% comp. | AEB | CV% comp. | AEB | CV% comp. |
| 200 | 0.8360 | 7% | 0.7042 | 12% | 1.0470 | 16% | 0.7481 | 8% | 0.7647 | 6% |
| 150 | 0.6185 | 7% | 0.5497 | 8% | 0.7831 | 17% | 0.5882 | 4% | 0.5689 | 6% |
| 100 | 0.4337 | 9% | 0.3752 | 10% | 0.5382 | 15% | 0.3729 | 11% | 0.4064 | 5% |
| 80 | 0.3400 | 7% | 0.2831 | 13% | 0.4256 | 16% | 0.3012 | 9% | 0.3000 | 9% |
| 50 | 0.2182 | 5% | 0.1848 | 12% | 0.2682 | 15% | 0.2003 | 6% | 0.1957 | 8% |
| 25 | 0.1117 | 4% | 0.0992 | 8% | 0.1381 | 15% | 0.1009 | 7% | 0.1101 | 1% |
| 10 | 0.0453 | 6% | 0.0421 | 5% | 0.0597 | 19% | 0.0458 | 1% | 0.0432 | 3% |
| 5 | 0.0255 | 13% | 0.0212 | 13% | 0.0287 | 8% | 0.0205 | 15% | 0.0236 | 5% |
| 1 | 0.0058 | 12% | 0.0054 | 5% | 0.0083 | 25% | 0.0062 | 5% | 0.0073 | 16% |
| 0 | 0.0022 | 22% | 0.0020 | 7% | 0.0026 | 11% | 0.0021 | 3% | 0.0023 | 1% |

| Calibration curve | Test 1 | Test 2 | Test 3 | Test 4 | Test 5 | Test 6 | Test 7 |
|---|---|---|---|---|---|---|---|
| pg/ml | DFT % | DFT % | DFT % | DFT % | DFT % | DFT % | DFT % |
| 200 | -4.48 | -6.03 | -1.56 | -1.07 | -0.32 | -0.05 | -1.85 |
| 150 | 1.44 | 2.77 | 1.23 | -2.12 | 0.82 | -3.53 | 0.38 |
| 100 | -0.95 | 0.16 | 0.34 | -1.35 | -1.23 | 3.40 | -5.00 |
| 80 | 3.60 | 4.65 | 0.91 | 5.86 | 0.51 | 3.15 | 4.57 |
| 50 | 4.43 | 1.58 | -1.30 | 3.23 | 0.78 | -1.78 | 2.67 |
| 25 | 2.17 | -1.64 | -0.12 | -2.33 | -0.33 | -0.48 | -5.80 |
| 10 | -2.88 | 1.03 | -0.78 | -5.98 | -4.75 | -10.31 | 3.83 |
| 5 | -23.87 | -14.18 | 4.63 | -2.05 | 5.25 | 7.46 | 2.53 |
| 1 | 17.80 | 11.03 | -3.91 | 6.68 | -2.64 | -1.79 | -6.40 |
| 0 | NaN | NaN | NaN | NaN | NaN | NaN | NaN |

**Supplementary table 1 and 2) Standard curve CV% and DFT% assessment.**

CV% calculated from average AEB values from 3 initial standard curves. Total of 7 assays carried out by 2 independent analysts. DFT = difference from total % predicted concentration of standards (pg/ml) versus actual.

| | Mean for 3 set of QCs on | | Comp. TEST4 to average | | Comp. TEST5 to average | | Comp. TEST6 to average | | Comp. TEST7 to average | |
|---|---|---|---|---|---|---|---|---|---|---|
| QCs | Mean AEB | CV% | Mean AEB | CV% comp. | Mean AEB | CV% comp. | Mean AEB | CV% comp. | Mean AEB | CV% comp. |
| HQC 140pg/ml | 0.5540 | 14% | 0.4940 | 8% | 0.7309 | 19% | 0.5160 | 5% | 0.4856 | 9% |
| MQC 75pg/ml | 0.3066 | 8% | 0.2714 | 9% | 0.4197 | 22% | 0.2752 | 8% | 0.3062 | 0% |
| LQC 15pg/ml | 0.0610 | 6% | 0.0526 | 10% | 0.0888 | 26% | 0.0568 | 5% | 0.0651 | 5% |

**Supplementary table 3) Assessment of quality control samples (QCs) CV%.**

| | Test 1 | Test 2 | Test 3 | Test 4 | Test 5 | Test 6 | Test 7 |
|---|---|---|---|---|---|---|---|
| QCs | DFT % | DFT % | DFT % | DFT % | DFT % | DFT % | DFT % |
| HQC 140pg/ml | 0% | 9% | 12% | 3% | 1% | 3% | 10% |
| MQC 75pg/ml | 7% | 9% | 6% | 4% | -5% | 22% | -4% |
| LQC 15pg/ml | 8% | 10% | 9% | 11% | -6% | 8% | 0% |

**Supplementary table 4) Assessment of quality control samples (QCs) DFT%.**

| Intraplate variability | Position 1 | | Position 2 | | Position 3 | | Average of 3 sets | |
|---|---|---|---|---|---|---|---|---|
| QCs | Mean AEB | CV% | Mean AEB | CV% | Mean AEB | CV% | Mean AEB | Total CV% |
| HQC 140pg/ml | 0.6394 | 6% | 0.6058 | 1% | 0.6149 | 2% | 0.6201 | 3% |
| MQC 75pg/ml | 0.3342 | 2% | 0.3305 | 0% | 0.3125 | 2% | 0.3257 | 4% |
| LQC 15pg/ml | 0.0651 | 1% | 0.0605 | 3% | 0.0618 | 1% | 0.0625 | 4% |

**Supplementary table 5) Intraplate variability assessment of CV%.**

| Preparation variability | Analyst 1 | | | | | | | | Analyst 2 | | | | | | | | | |
|---|---|---|---|---|---|---|---|---|---|---|---|---|---|---|---|---|---|---|
| | Prep. 1 | | Prep. 2 | | Prep. 3 | | Assay Mean | | Prep. 1 | | Prep. 2 | | Prep. 3 | | Assay Mean | | Total Mean | Total CV |
| QCs | Mean AEB | CV% | Mean AEB | CV% | Mean AEB | CV% | Mean AEB | CV% | Mean AEB | CV% | Mean AEB | CV% | Mean AEB | CV% | Mean AEB | CV% | Mean AEB | CV% |
| HQC 140pg/ml | 0.5160 | 1% | 0.5140 | 3% | 0.5180 | 2% | 0.5160 | 0% | 0.4940 | 1% | 0.4814 | 2% | 0.4675 | 2% | 0.4930 | 2% | 0.4985 | 4% |
| MQC 75pg/ml | 0.2752 | NaN | 0.2835 | 3% | 0.2852 | 9% | 0.2813 | 2% | 0.2714 | 1% | 0.2831 | 2% | 0.2666 | 6% | 0.2876 | 0% | 0.2775 | 3% |
| LQC 15pg/ml | 0.0568 | 3% | NaN | 14% | 0.0604 | 2% | 0.0586 | 4% | 0.0526 | 3% | 0.0540 | 12% | 0.0526 | 5% | 0.0594 | 11% | 0.0553 | 6% |

**Supplementary table 6) Reproducibility assessment using independently prepared QCs.**

| Matrix control: QC4 | Test 1 | Test 2 | Test 3 | Test 4 | CV% |
|---|---|---|---|---|---|
| Mean AEB | 0.0526 | 0.0576 | 0.0696 | 0.0553 | 13% |
| Predicted concentration pg/ml | 26.7 | 28.1 | 24.6 | 25.2 | 6% |

**Supplementary table 7) Reproducibility of matrix control CV%.**

| | **neat - anchore** | 1:2 | | | 1:4 | | | 1:8 | | | 1:16 | | |
|---|---|---|---|---|---|---|---|---|---|---|---|---|---|
| | **Mean conc.** | **Mean conc.** | **Predicted neat** | Error% | **Mean conc.** | **Predicted neat** | Error% | **Mean conc.** | **Predicted neat** | Error% | **Mean conc.** | **Predicted neat** | Error% |
| **S1** | 15.16 | 13.76 | 27.5 | -82% | 7.40 | 29.6 | -95% | 4.39 | 35.2 | -132% | 2.36 | 37.7 | -149% |
| **S2** | 11.75 | 9.96 | 19.9 | -70% | 5.66 | 22.6 | -92% | 3.19 | 25.5 | -117% | 1.37 | 21.8 | -86% |
| **S3** | 11.68 | 9.58 | 19.2 | -64% | 6.68 | 26.7 | -129% | 3.21 | 25.7 | -120% | 1.59 | 38.8 | -232% |
| **S4** | 21.84 | 17.63 | 35.3 | -61% | 11.68 | 46.7 | -114% | 6.16 | 49.3 | -126% | 2.70 | 43.2 | -98% |
| **S5** | 11.68 | 9.08 | 18.2 | -55% | 5.78 | 23.1 | -98% | 3.22 | 25.8 | -121% | 1.03 | 25.3 | -117% |
| **S6** | 8.62 | 9.41 | 18.8 | -118% | 5.67 | 22.7 | -163% | 3.19 | 25.5 | -196% | 1.21 | 29.6 | -243% |
| **Average:** | | | | -75% | | | -115% | | | -135% | | | -154% |

| | **1:2 - anchore** | 1:4 | | | 1:8 | | | 1:16 | | |
|---|---|---|---|---|---|---|---|---|---|---|
| | **Mean conc.** | **Mean conc.** | **Predicted 1:2** | Error% | **Mean conc.** | **Predicted 1:2** | Error% | **Mean conc.** | **Predicted 1:2** | Error% |
| **S1** | 13.76 | 7.40 | 14.8 | 7% | 4.39 | 17.6 | -28% | 2.36 | 18.9 | -37% |
| **S2** | 9.96 | 5.66 | 11.3 | -14% | 3.19 | 12.8 | -28% | 1.37 | 10.9 | -10% |
| **S3** | 9.58 | 6.68 | 13.4 | -39% | 3.21 | 12.8 | -34% | 1.59 | 19.4 | -102% |
| **S4** | 17.63 | 11.68 | 23.4 | -32% | 6.16 | 24.6 | -40% | 2.70 | 21.6 | -22% |
| **S5** | 9.08 | 5.78 | 11.6 | -27% | 3.22 | 12.9 | -42% | 1.03 | 12.6 | -39% |
| **S6** | 9.41 | 5.67 | 11.3 | -21% | 3.19 | 12.8 | -36% | 1.21 | 14.8 | -57% |
| **Average:** | | | | -21% | | | -35% | | | -45% |

**Supplementary table 8)** Dilutional parallelism was assessed by running CSF from six C9orf72 expansion positive donors either neat, 1:2, 1:4, 1:8 and 1:16 in diluent A**.**

| QC4 | Meas. 1 | Meas. 2 | Mean AEB | CV% | Pred. conc. (pg/ml) | CV% for mean AEB | CV% for pred. conc (pg/ml) |
|---|---|---|---|---|---|---|---|
| Fresh | 0.0737 | 0.0654 | 0.0696 | 8% | 24.6 | 4% | 5% |
| Freeze-thaw 1 | 0.0805 | 0.0691 | 0.0748 | 11% | 26.5 | | |
| Freeze-thaw 2 | 0.0732 | 0.0769 | 0.0751 | 4% | 26.6 | | |
| Freeze-thaw 3 | 0.0698 | 0.0685 | 0.0691 | 1% | 24.4 | | |

**Supplementary table 9) Freeze-thaw stability of poly(GP) from human CSF.**

| pg/ml | Mean AEB | | | | CV% fresh vs. F-T 1 | CV% fresh vs. F-T 2 | CV% fresh vs. F-T 3 | % Error in prediction concentration | | | |
|---|---|---|---|---|---|---|---|---|---|---|---|
| | Fresh | Freeze-thaw 1 | Freeze-thaw 2 | Freeze-thaw 3 | | | | Fresh | Freeze-thaw 1 | Freeze-thaw 2 | Freeze-thaw 3 |
| 200 | 0.7481 | 0.7675 | 0.7258 | 0.7303 | 2% | 2% | 2% | -0.05 | -1.22 | -3.51 | -11.97 |
| 150 | 0.5882 | 0.5773 | 0.5568 | 0.5373 | 1% | 4% | 6% | -3.53 | 1.43 | -0.01 | -3.75 |
| 100 | 0.3729 | 0.3964 | 0.3822 | 0.3662 | 4% | 2% | 1% | 3.40 | 1.42 | 2.00 | -0.74 |
| 80 | 0.3012 | 0.3280 | 0.3003 | 0.3001 | 6% | 0% | 0% | 3.15 | -0.75 | 5.55 | -1.05 |
| 50 | 0.2003 | 0.2067 | 0.2086 | 0.1846 | 2% | 3% | 6% | -1.78 | 0.89 | -3.28 | 4.52 |
| 25 | 0.1009 | 0.1167 | 0.1004 | 0.0978 | 10% | 0% | 2% | -0.48 | -9.09 | 1.52 | 2.87 |
| 10 | 0.0458 | 0.0426 | 0.0456 | 0.0375 | 5% | 0% | 14% | -10.31 | 5.27 | -11.82 | 12.17 |
| 5 | 0.0205 | 0.0231 | 0.0200 | 0.0215 | 8% | 2% | 3% | 7.46 | 2.23 | 4.53 | 3.90 |
| 1 | 0.0062 | 0.0064 | 0.0054 | 0.0072 | 2% | 10% | 10% | -1.79 | -3.95 | 2.52 | -34.34 |
| 0 | 0.0021 | 0.0018 | 0.0020 | 0.0013 | 14% | 4% | 33% | NaN | NaN | NaN | NaN |

**Supplementary table 10) Freeze-thaw stability of GST-GP32 standard.**
